# Bounding the levels of transmissibility & immune evasion of the Omicron variant in South Africa

**DOI:** 10.1101/2021.12.19.21268038

**Authors:** Carl A. B. Pearson, Sheetal P. Silal, Michael W.Z. Li, Jonathan Dushoff, Benjamin M. Bolker, Sam Abbott, Cari van Schalkwyk, Nicholas G. Davies, Rosanna C. Barnard, W. John Edmunds, Jeremy Bingham, Gesine Meyer-Rath, Lise Jamieson, Allison Glass, Nicole Wolter, Nevashan Govender, Wendy S. Stevens, Lesley Scott, Koleka Mlisana, Harry Moultrie, Juliet R. C. Pulliam

## Abstract

A new SARS-CoV-2 variant of concern, Omicron (B.1.1.529), has been identified based on genomic sequencing and epidemiological data in South Africa. Presumptive Omicron cases in South Africa have grown extremely rapidly, despite high prior exposure and moderate vaccination coverage. The available evidence suggests that Omicron spread is at least in part due to evasion of this immune protection, though Omicron may also exhibit higher intrinsic transmissibility. Using detailed laboratory and epidemiological data from South Africa, we estimate the constraints on these two characteristics of the new variant and their relationship. Our estimates and associated uncertainties provide essential information to inform projection and scenario modeling analyses, which are crucial planning tools for governments around the world.

**One Sentence Summary:** We report a region of plausibility for the relative transmissibility and immune escape characteristics of the SARS-CoV-2 Omicron variant estimated by integrating laboratory and epidemiological data from South Africa.

## Background

As of 6^th^ December 2021, over 265M SARS-CoV-2 infections have been reported worldwide, including 5.2M deaths. Approximately 87% of those cases and 76% of deaths have occurred since the first variants of concern were detected in December 2020 (1–3), despite increasing coverage with efficacious vaccines, in particular in high-income countries. In November 2021, South African researchers reported a new variant designated B.1.1.529 (4), subsequently declared variant of concern Omicron by the World Health Organization (WHO) on November 26^th^ (5). Genomic sequencing of Omicron indicates a large number of mutations in the receptor binding domain of the spike protein (6), and prior work has shown that such mutations lead to decreases in virus neutralization (7, 8). Ongoing neutralization experiments consistently find reductions in neutralization of Omicron, though the data available to date indicate a wide range for the magnitude of these reductions (9–12). Furthermore, ongoing surveillance monitoring for changes in reinfection risk in South Africa finds an early signal of immune evasion at the population level that coincides temporally with the emergence of the Omicron variant (13). Critically, a major sublineage of the Omicron variant, designated BA.1, which has come to dominate transmission in South Africa, is characterized by a deletion at sites 69-70 of the spike protein (4). The same change was previously found to cause S-gene target failure (SGTF) for the Alpha variant in the TaqPath™ COVID-19 (Thermo Fisher Scientific, Waltham, MA, USA) PCR test (14, 15), and researchers have confirmed that this deletion in Omicron also leads to SGTF (16).

To perform public health model-based scenario analysis of Omicron impact, researchers need estimates of the transmission characteristics of the new variant, particularly the degrees of change in intrinsic transmissibility and immune evasion. Using epidemiological and SGTF time-series data and prior model inferences, we report a Next-Generation-Matrix (NGM) based-approach to estimate a constraint curve for these two properties that is compatible with observed Omicron case growth in South Africa. We incorporate estimates of protection against infection based on recent neutralization experiments to narrow the plausible range of immune evasion.

## Methods in brief

For four of South Africa’s nine provinces, using positive SARS-CoV-2 testing data with specimen receipt dates from October 1^st^ through December 6^th^ 2021, we performed the following analysis:

1. Fit the proportion-SGTF time series to estimate the relative growth rate (Δr) of proportion Omicron sublineage BA.1 (hereafter, BA.1) versus other circulating lineages (hereafter, background; presumed to be primarily Delta, but with a slow-growing C.1.2 fraction (*17*)).
2. Using profile confidence interval of the BA.1 proportion estimates, create samples of paired BA.1 and background infection series from the time series of detected infections (i.e., primary infections plus reinfections).
3. Estimate paired daily time-varying reproduction number (R_t_) series for BA.1 and background infections, assuming either a generation time of 6.4 days for both BA.1 and background, or a shorter generation time of 5.2 days for BA.1. Compute the daily ratios of *R*_*t*_ values.
4. Take the geometric mean of each ratio ensemble for the period 14^th^ - 20^th^ November to create an estimated distribution of *R*_*t*_ ratios.
5. Varying the level of immune evasion for BA.1, compute next generation matrix principal eigenvalue (NGM R) ratios.
6. Identify the required transmissibility multiplier distribution to match the *R*_*t*_ ratios from step 4 to the principal eigenvalue ratios in step 5.

In a sensitivity analysis, we repeated these analyses using a cut-off date of 27^th^ November, before testing increased as a result of the announcement of a new variant. We performed all analyses and visualization using R 4.1.2, using the following packages: data.table (*18*), EpiNow2 (*19*), ggplot2 (*20*), patchwork (*21*), bbmle (*22*), and emdbook (*23*).

### Relative Growth Estimation & Incidence Ensembles

We used province-specific time-series counts (adjusted to correct for reporting delays as in (*13*)) of positive SARS-CoV-2 PCR tests performed by South Africa’s National Health Laboratory Service (NHLS) and Lancet Laboratories, a private sector laboratory. Both laboratories use the TaqPath™ COVID-19 assay, which exhibits the SGTF associated with BA.1. Laboratory data included gene specific cycle threshold (Ct) values for positive PCR tests, and only samples with Ct values ≤30 on at least one gene target (ORF1ab or nucleocapsid) were included in the growth rate analysis.

We used maximum likelihood to fit a separate model for each province, explaining the observed data with logistic growth and imperfect test performance. We fit a logistic growth rate, half-replacement time, and SGTF sensitivity (the proportion of BA.1 infections with SGTF) and specificity (fraction of positive tests for which the assay properly binds the S-gene in background infections). We attempted to fit each province with a beta-binomial distribution to account for over-dispersion, as well as with a binomial distribution, and calculate the associated profile confidence intervals (*23*). We rejected fits that did not converge, fits where the confidence interval for Δr did not converge, and (for the beta-binomial) fits where the shape parameter exceeded a set threshold of exp(3) ∼ 20 (meaning that the overdispersion was not supported). When neither model was rejected, we used the beta-binomial model. We report results for provinces where at least one model converged (Gauteng (GP), KwaZulu-Natal (KZN), Northern Cape (NC), and Northwest (NW)).

We then generated ensembles of BA.1 proportion time series from the fits using posterior predictive sampling (*23*). We use those time series to create 1,000 sample partitions of the observed incidence into BA.1 and background infections.

### Aggregate R_*t*_ Estimation

Using the paired BA.1 and background infection time series samples, we estimated the R_t_ value for each series assuming a gamma distributed intrinsic generation interval with mean of 6.38 days (s.d. of mean: 1.24; s.d. 5.39, s.d. of s.d. 1.35) (*24*). As a sensitivity analysis, we also estimate the R_t_ ratio with a generation interval with a mean of 5.2 days for the BA.1 infection time series, which we implement by halving the value of the incubation period. We create a posterior sample of 1,000 R_*t*_ time series estimates for each province. Matching on sample pairs, we compute the series of R_*t*_ ratios for BA.1 to background. Finally, for each sample we compute a geometric mean ratio for the week of 14^th^ to 20^th^ November (inclusive). This period is after the initial noisy introduction of BA.1 but prior to the surge in testing associated with the public announcement of the variant’s discovery (see Supplement Fig S2). After taking the geometric mean, we recover a distribution of BA.1 to background time-averaged R_t_ ratios during a period of relatively stable growth.

### Mobility & Contact Matrices

We adapted a global set of national contact matrices (*25*) by adjusting simultaneously for symmetry of contacts and provincial age distributions to create province-specific matrices. As in other work (*26*), we computed the weighted geometric, centered 7-day average of Google Mobility indicators, consolidating the “other” contacts multiplier by weighting retail & recreation 0.3, grocery & pharmacy 0.3, transit 0.3, and parks 0.1. We use the Oxford Stringency index (*27*) for schools weighting, using the C1 (Containment and closure policies: School closing) value * ⅓.

### Non-Susceptible Fractions

The immune-escapable (recovered and/or vaccinated) and currently infected fractions of the population were estimated using the South African National COVID-19 Epi Model (NCEM). NCEM is a mathematical transmission model developed by the South African COVID-19 Modelling Consortium (SACMC) that has been adapted several times over the course of the COVID-19 pandemic (*28*). The current version (v6.0) is a multi-strain, age-structured, spatially-explicit, generalized susceptible-exposed-infected-recovered (SEIR) compartmental model of COVID-19 that runs at the scale of South Africa’s provinces (*29*). The model, which accounts for South Africa’s SARS-CoV-2 vaccination rollout by age-group, province, and vaccine doses and product type, has been calibrated to provincial and age-specific data on seroprevalence, hospital admissions, SARS-CoV-2 variant distribution, and hospital-based and excess mortality. The current version of the model incorporates the dynamics of the ancestral, Beta, and Delta variants, each of which drove a major prior wave of SARS-CoV-2 transmission in South Africa.

We use the model to provide a time series, by age and province, of the proportion of the population protected from infection by vaccines, by prior infection, and by current infection. The model assumes that vaccine-derived protection from infection wanes on average six months post-vaccination, while protection against severe disease is retained over the modelled time frame (*30*–*32*).

### Next Generation Matrix Method

The Next-Generation-Matrix (NGM) method (*33*) can be used to compute the basic reproduction number (R_0_) for a compartmental model of a pathogen, and given the fraction of the population in infectable classes, an effective reproduction number can also be computed. We assume an age-stratified population, with susceptible, exposed, infectious, recovered, and vaccinated compartments, with additional distinctions within the infectious compartment into asymptomatic, pre-symptomatic, and symptomatic compartments. We assume age-specific susceptibility and symptomatic fractions (*34*).

For the estimation period, we also used estimates from the NCEM to set the susceptible proportion. We then increased the susceptible proportion uniformly across all ages by a specified multiplicative factor to represent different scenarios of immune evasion.

### Estimating the Plausible Range for Immune Escape

Using an analysis that synthesizes recent neutralization results with prior experiments to correlate vaccine efficacy against infection (*35, 36*), we can infer the relative reduction in protection due to BA.1 versus Delta, which we presume is the primary circulating variant in the background transmission.

## Results

We found that BA.1 has a growth advantage over the background mixture of SARS-CoV-2 lineages circulating in October and November 2021. This advantage appears in the time series of the proportion SGTF for all provinces in South Africa. However, the maximum likelihood estimation procedure does not converge for every province, likely reflecting the heterogenous geographic distribution of laboratories with capacity to perform the TaqPath assay. In the four provinces for which the model fits did converge (Fig 1, Table 1), the transformation of the infection time series demonstrated bi-exponential trends, which indicates continuing decline of background lineages and rapid growth of BA.1 (Fig 2). The estimated relative multipliers for those trends (Fig 3) indicate a larger growth advantage for BA.1 than observed with any prior variant emergence event. When combined with estimates of historical accumulation of immune protection due to vaccination and/or infection, the large multipliers suggest that BA.1 has an intrinsic transmission advantage (Fig 4). This conclusion is robust for most of our sensitivity analyses, however if the generation interval is shorter than for previously circulating variants or the true susceptible proportion is lower than we have estimated, reduced intrinsic transmissibility is plausible (Fig 5).

**Table 1.**
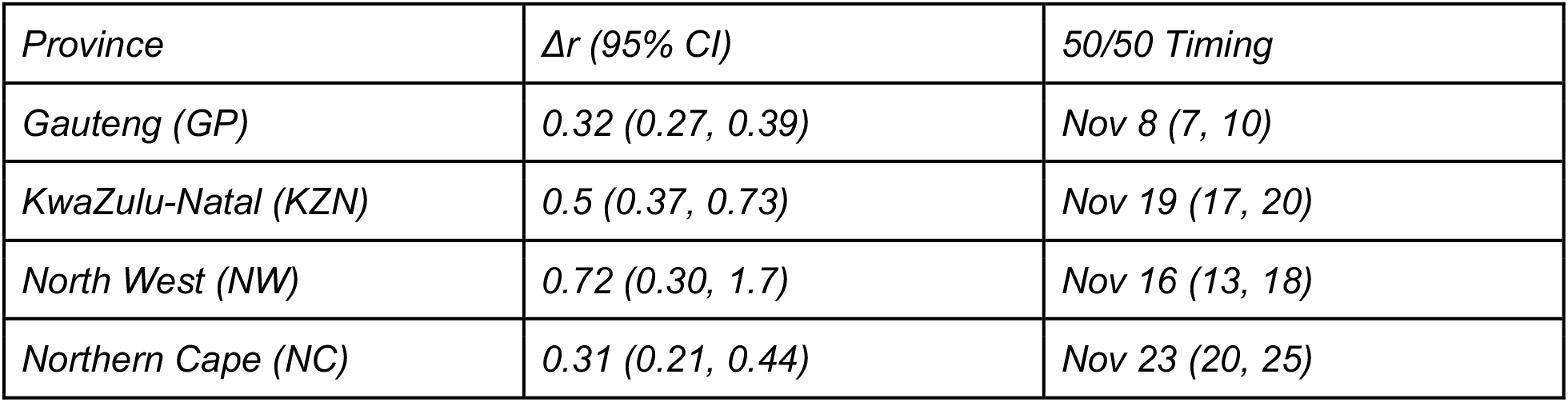
Δr values by model and province. *Δr* is the growth advantage of Omicron sublineage BA.1 relative to background variants. *50/50 Timing* is an estimate of the date on which half of all samples were BA.1.

| Province | $\Delta r$ (95% CI) | 50/50 Timing |
| --- | --- | --- |
| Gauteng (GP) | 0.32 (0.27, 0.39) | Nov 8 (7, 10) |
| KwaZulu-Natal (KZN) | 0.5 (0.37, 0.73) | Nov 19 (17, 20) |
| North West (NW) | 0.72 (0.30, 1.7) | Nov 16 (13, 18) |
| Northern Cape (NC) | 0.31 (0.21, 0.44) | Nov 23 (20, 25) |

**Fig 1:**
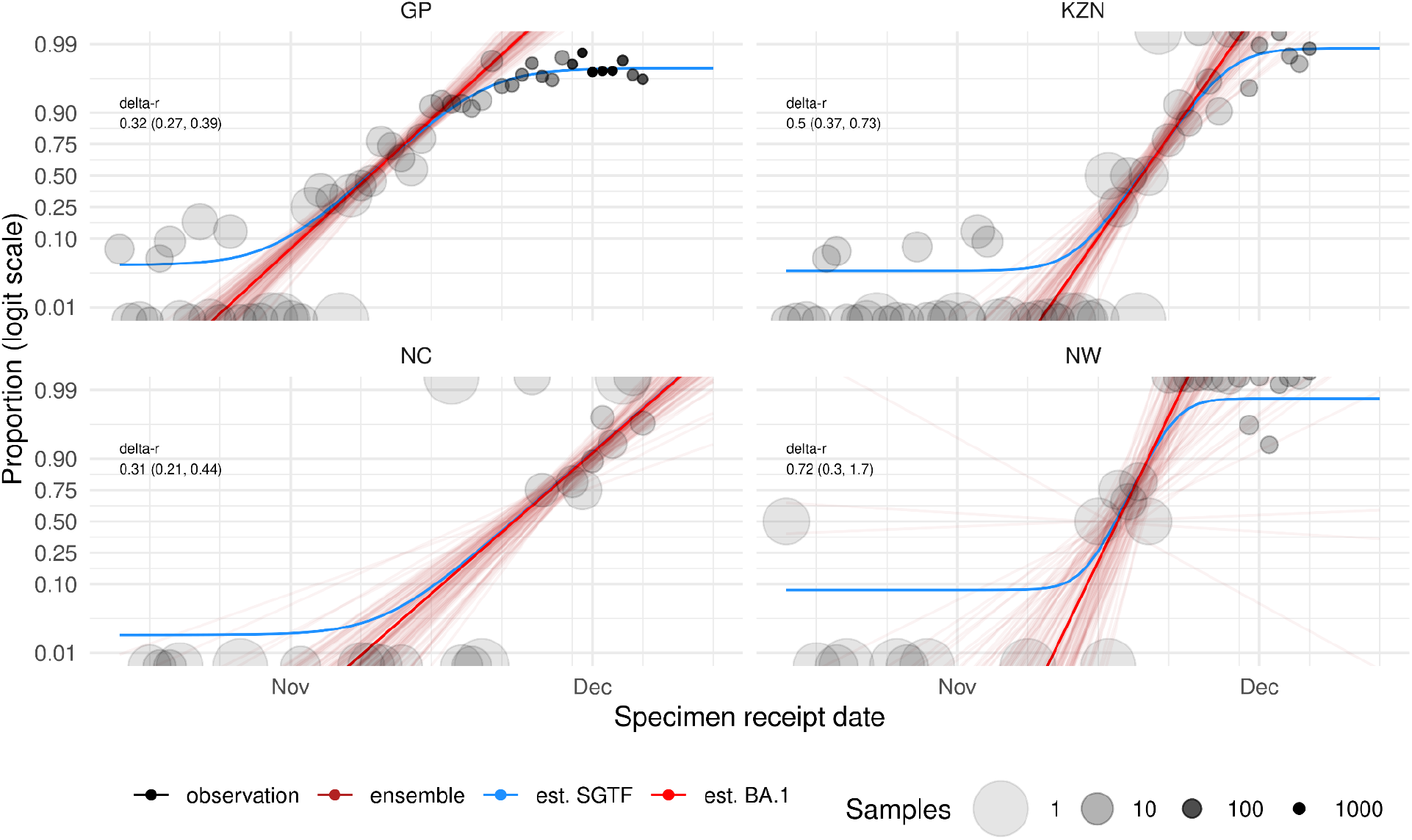
Estimation of the growth advantage of BA.1 based on S-gene target failure (SGTF) proportion through time. Circles represent the observed proportion of SGTF by sample receipt date and province (the area of each circle is inversely related to number of samples, so that larger areas correspond to more uncertainty). Light red lines show simulated time series of BA.1 prevalence using 50 parameter sets from posterior predictive sampling, in each province for which we have an acceptable model fit. We also show the maximum-likelihood estimates for growth rate and timing (solid red line), as well as the overall fitted SGTF (blue line), which incorporates sensitivity and specificity estimates. The estimates shown here are generated using model fits that assume a beta-binomial error distribution and account for sensitivity and specificity of the SGTF as a marker of BA.1. GP: Gauteng; KZN: KwaZulu-Natal; NC: Northern Cape; NW: North West.

**Fig 2:**
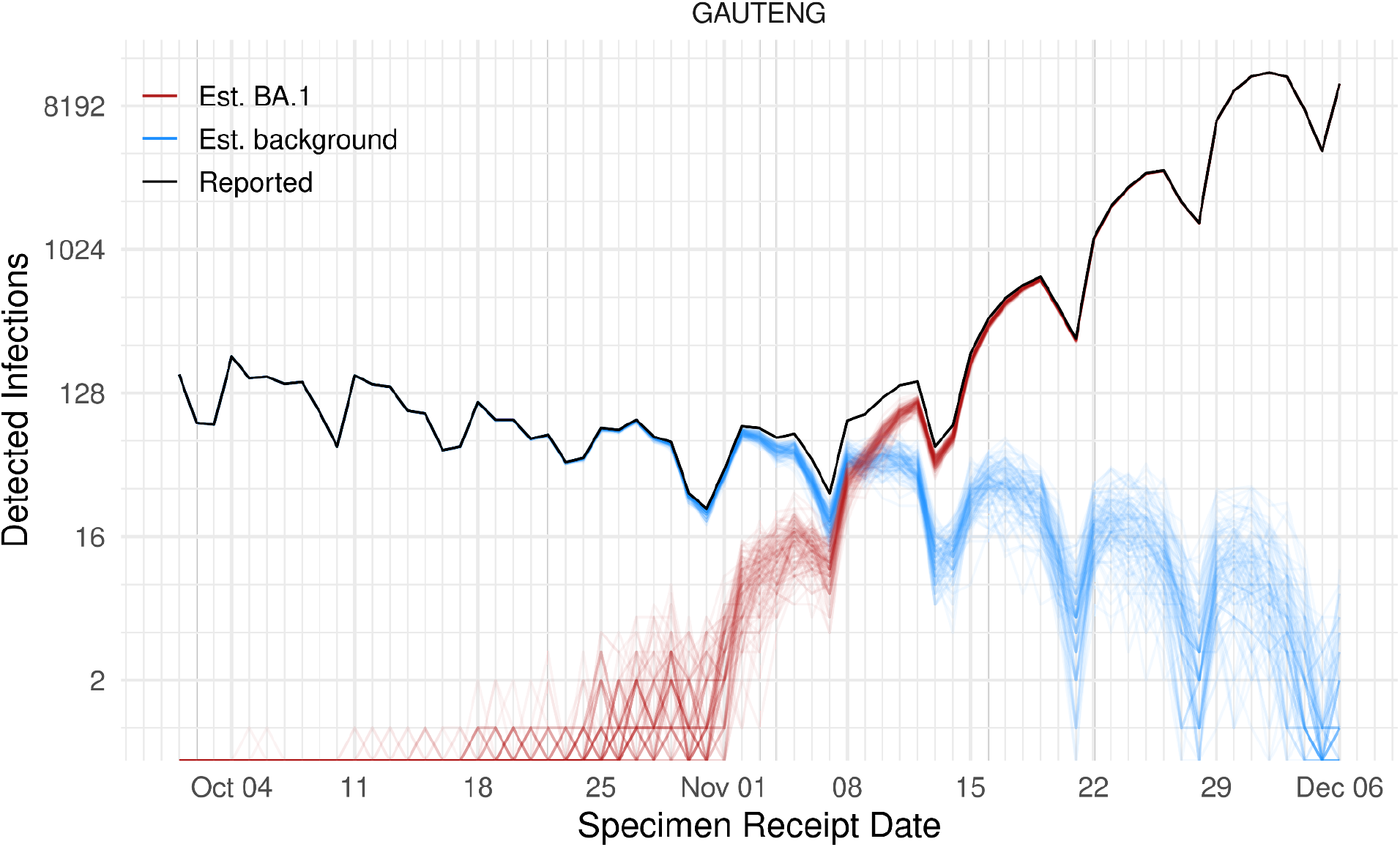
Incidence of detected infection split into BA.1 and background in Gauteng province. Applying the estimated BA.1 versus background fractions shown in Fig 1, we sample potential infection trajectories, which are then fed into the R_t_ estimation stage. The black curve shows detected infections by sample receipt date, with red and blue curves illustrating sampled BA.1 and background infection trajectories, respectively.

**Fig 3:**
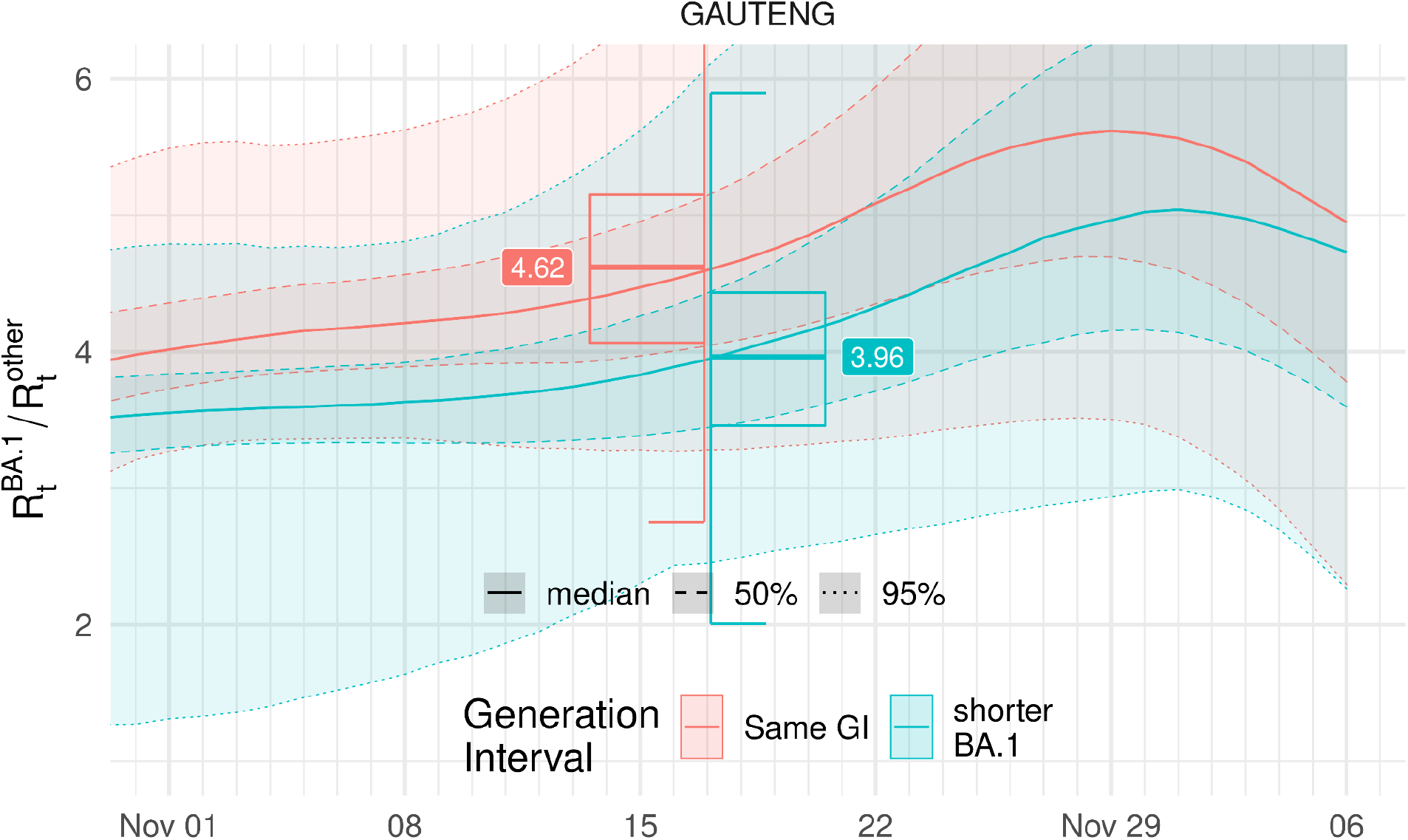
Ratio of time-varying reproduction numbers in Gauteng province. Estimated ratio of BA.1 R_t_ to background variant R_t_, during the emergence period of BA.1 in Gauteng province, South Africa. We aggregate the time after super-spreading events in early November and before the announcement of Omicron sublineage BA.1, taking the geometric mean for the period 14^th^ - 20^th^ November 2021. Box plots show the median, interquartile and 95% ranges of the time-averaged R_t_ ratio, for the main parameter set (red) and for a sensitivity analysis that considers the possibility of a shorter incubation period (and therefore generation interval) for BA.1 (cyan).

**Fig 4:**
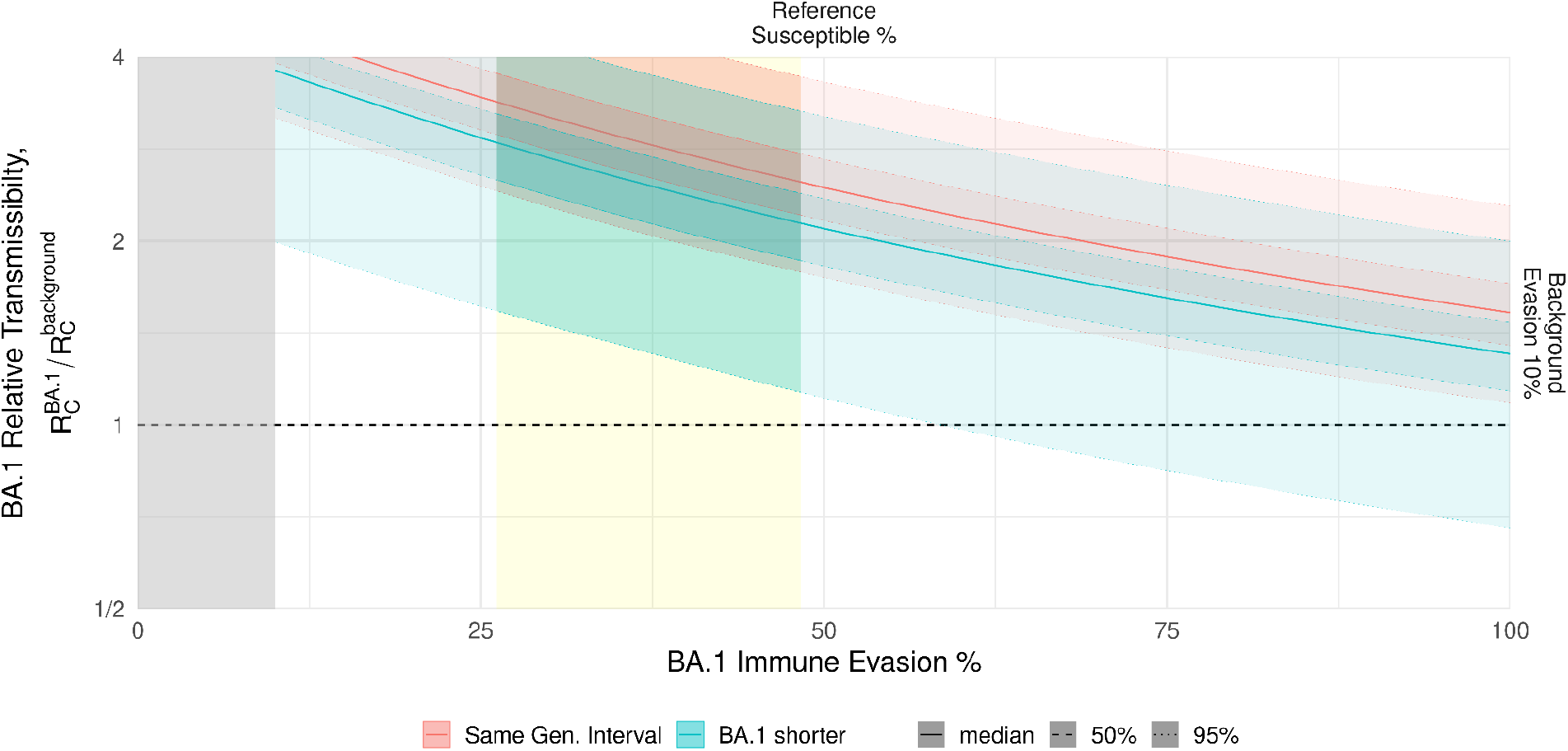
Estimated transmissibility & immune evasion relationship for reference scenario based on estimates for Gauteng province, South Africa. The red and cyan shaded regions indicate the region of plausibility for relative transmissibility and immune evasion values for BA.1, assuming no change in the generation interval or a shorter generation interval, respectively. The yellow band represents estimated plausible immune escape values based on preliminary neutralization data *(36)* and previously estimated relationships between neutralization fold-reduction and vaccine efficacy against infection *(37)*. The grey band represents values of immune protection that are considered implausible because they would imply greater levels of immune evasion for background variants than for BA.1. The horizontal dashed line indicates equal transmissibility for BA.1 and background variants.

**Fig 5:**
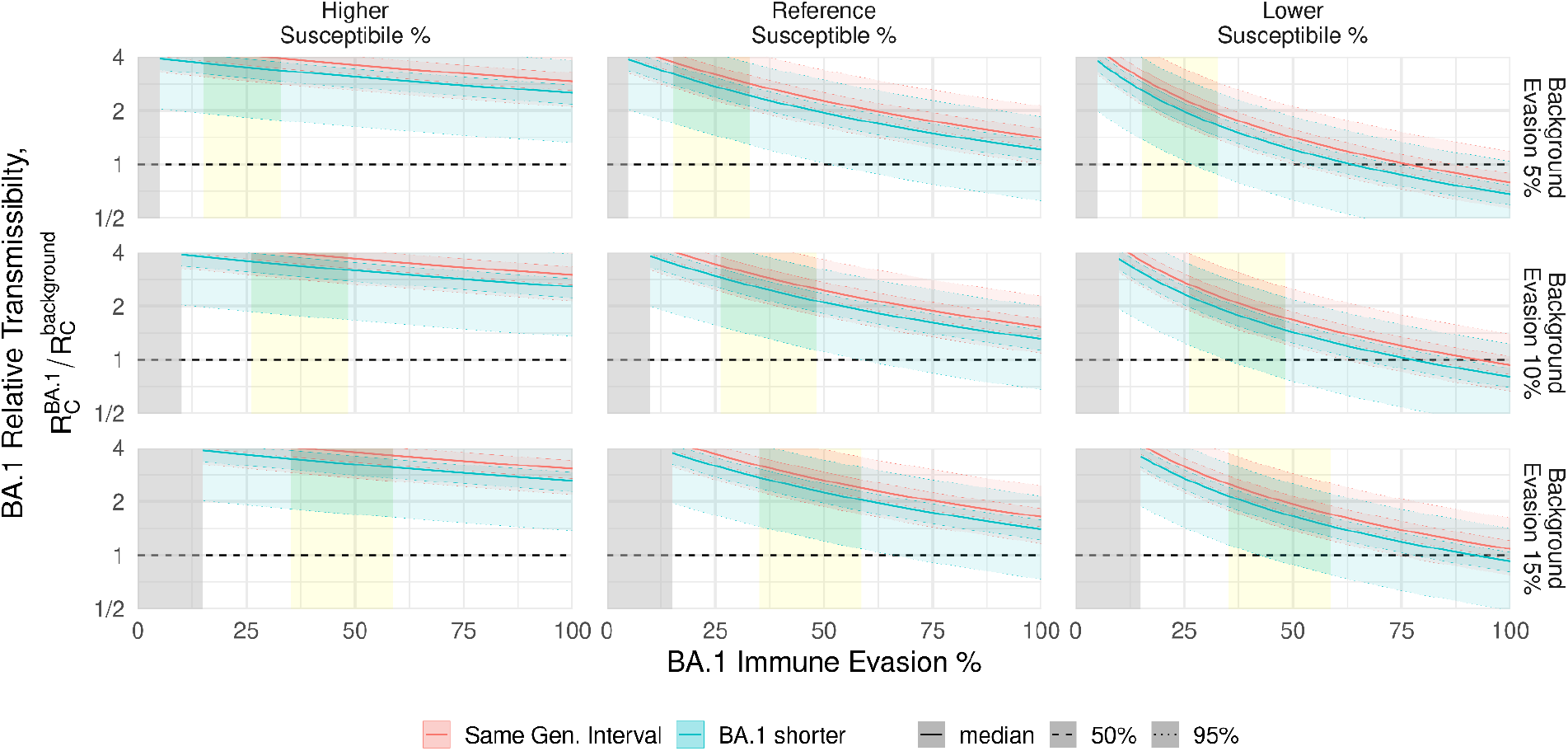
Sensitivity of the plausible estimated transmissibility & immune evasion relationship. These panels compute the same calculation as shown in Fig 4, but with varying assumptions regarding the underlying fully susceptible proportion (columns) and the background level of protection provided by prior infection (rows). The main analysis, as shown in Fig 4, is replicated in the center panel.

While laboratory experiments find a consistent fold-decrease in antibody neutralization of Omicron virus and pseudovirus, the reference starting neutralization varies extensively based on the kind of immune protection, i.e. from which vaccine product(s) and dosing regimen, time from latest vaccination, and whether the individual acquired additional immune protection from a prior infection (*9, 10, 12, 36*). In our modelling framework, we assume that the population mixes homogeneously, and the appropriate aggregation of these immune states is not clear. As such, we consider variations on the assumed level of protection by considering alternative amounts of background immune evasion and fractions with some protection and framing the level of immune evasion by BA.1 relative to this background (Fig 5). For our main analysis, this approach suggests a 3- to 8-fold reduction in neutralization titres for Omicron relative to Delta, corresponding to immune evasion of 26 - 48% (15 - 33% for low Delta evasion scenario, 35 - 59% for high Delta evasion).

## Discussion

The Omicron variant, and specifically sublineage BA.1, is outcompeting the background mixture of SARS-CoV-2 variants in the South African setting and is doing so in the presence of high population-level immunity. Early indications from around the globe suggest a similar growth advantage in other settings. Our findings imply that BA.1’s competitive advantage most likely results from a combination of higher intrinsic transmissibility and an ability to evade immunity from prior infection and/or vaccination. Although the precise values for transmissibility and immune evasion are not identifiable in this analysis, the plausible ranges may be narrowed further when our results are combined with data from additional neutralization studies and/or population-level observational studies of the protective efficacy provided by vaccination or prior infection.

For most settings, the most useful comparison would be the relative transmissibility of BA.1 to Delta, which remains the predominant variant circulating in most countries. The background mixture of variants in South Africa appears to be predominantly Delta during the time we are comparing growth rates (*38*).

Our results imply that Omicron is likely more transmissible than Delta, even at extreme levels of immune evasion, though some of the alternative assumptions we considered can be compatible with lower transmissibility in a fully susceptible (“immunologically naive”) population. Estimates for the relative transmissibility of a new SARS-CoV-2 variant can vary substantially among regions within countries (*39*), and accordingly, the measured relative transmissibility of Omicron may differ in other settings.

Our NGM approach assumes that individuals with breakthrough infections are subsequently as infectious in terms of their onward transmission as infections in immunologically naive individuals; this approximation ultimately tends to underestimate the BA.1 transmission advantage over other circulating variants. We expect that prior immunity will reduce the severity of breakthrough infections, and prior immunity has previously been correlated with lower propensity for onward transmission. If individuals with prior immunity are less transmissible, the selective advantage of immune evasion may be reduced, suggesting that population models accounting for this effect would tend to find higher transmissibility multipliers for equivalent immune evasion levels to match the empirical trends. We also assume similar infectious periods for BA.1 compared to other variants. If in fact BA.1 has a faster generation time, e.g. due to a shorter infectious period, then the estimated transmissibility multiplier would be still higher, offsetting the slightly lower R_t_ ratio calculated when assuming shorter generation times in our sensitivity analysis.

Given the epidemic trends in South Africa, the rapid introduction of BA.1 to other regions, and broad remaining uncertainty, public health analysis and planning should pivot to consider the impact of the Omicron variant in other locales. Our results imply that Omicron infections are likely to become widespread in all contexts, and while severity estimates of Omicron are still developing (*40*), the foundational work to prepare national and regional response scenarios for hospitalization and deaths can begin immediately. Given the complex constellation of sequence changes in BA.1, broad genomic surveillance should continue, particularly to calibrate the use of indicators like SGTF. The scale of these genomic changes also suggests the potential for changes in clinical presentation of COVID-19, which should be evaluated to inform projection analyses. Wherever household studies or other epidemiological fieldwork and surveillance are available to pivot, extend, or restart, best efforts should be made to measure transmissibility, immune escape, and generation times more directly.

## Data Availability

The code for all the analyses will soon be available from https://github.com/SACEMA/omicronSA. Data necessary to conduct the analysis is either included in the repository, fetched by that code from publicly available sources, or available upon formal request. Aside from the data made available in the repository, the non-public South African data are covered by a non-disclosure agreement and cannot be released by the authors. Requests for these data must be made in writing to the National Institute for Communicable Diseases, South Africa.

## Acknowledgements

We thank colleagues in the SARS-CoV-2 Variants Research Consortium in South Africa for valuable discussion during the preparation of this work, and Sebastian Funk for rapid pre-submission peer review. We acknowledge the Network for Genomic Surveillance - South Africa (NGS-SA) led by Prof Tulio de Oliveira for its role in discovery of the Omicron variant and validation of the S-gene target failure as a marker of BA.1 sublineage. We also wish to acknowledge the members of the NICD Epidemiology and Information Technology teams that curate, clean, and prepare the data utilized in this analysis.

## Epidemiology team

Andronica Moipone Shonhiwa, Genevie Ntshoe, Joy Ebonwu, Lactatia Motsuku, Liliwe Shuping, Mazvita Muchengeti, Jackie Kleynhans, Gillian Hunt, Victor Odhiambo Olago, Husna Ismail, Nevashan Govender, Ann Mathews, Vivien Essel, Veerle Msimang, Tendesayi Kufa-Chakezha, Nkengafac Villyen Motaze, Natalie Mayet, Tebogo Mmaborwa Matjokotja, Mzimasi Neti, Tracy Arendse, Teresa Lamola, Itumeleng Matiea, Darren Muganhiri, Babongile Ndlovu, Khuliso Ravhuhali, Emelda Ramutshila, Salaminah Mhlanga, Akhona Mzoneli, Nimesh Naran, Trisha Whitbread, Mpho Moeti, Chidozie Iwu, Eva Mathatha, Fhatuwani Gavhi, Masingita Makamu, Matimba Makhubele, Simbulele Mdleleni, Bracha Chiger, Jackie Kleynhans

## Information Technology team

Tsumbedzo Mukange, Trevor Bell, Lincoln Darwin, Fazil McKenna, Ndivhuwo Munava, Muzammil Raza Bano, Themba Ngobeni

Finally, we would like to acknowledge the teams within the National Institute for Communicable Diseases Centre for Respiratory Diseases and Meningitis (Anne von Gottberg, Daniel Amoako, Josie Everatt, Linda de Gouveia, Thabo Mohale, Boitshoko Mahlangu, Noxolo Ntuli, Anele Mnguni, Gerald Motsatsi, Malusi Ndlovu, Noluthando Duma), Centre for HIV and STIs (Catherine Scheepers and Jinal Bhiman), and Sequencing Core Facility (Arshad Ismail, Annie Chan, Morne du Plessis). We thank all laboratories for submitting specimens for sequencing, and all public laboratories and Lancet Laboratories for Thermo Fisher TaqPath PCR data. We would further like to acknowledge Graeme Dor, Beth Crankshaw, Silence Ndlovu, Helena Vreede and Pedro da Silva for data analytics and oversight on monitoring the population and assay specific Ct value changes using the NHLS laboratory data.

## Data Availability

The code for all the analyses is available from https://github.com/SACEMA/omicronSA. Data necessary to conduct the analysis is either included in the repository, fetched by that code from publicly available sources, or available upon formal request. Aside from the data made available in the repository, the non-public South African data are covered by a non-disclosure agreement and cannot be released by the authors. Requests for these data must be made in writing to the National Institute for Communicable Diseases, South Africa.

## Supplementary Information

*Nota Bene in re Implied R0s for BA.1 & Delta*

The 100% immune evasion end of main text Fig 4 should be interpreted as estimating the *R*_*0*_ *under “control”* (i.e., after the response is underway), or R_c_, for Omicron sublineage BA.1, and by implication of the transmissibility multiplication factor, for the background lineage composition also under control conditions (more specifically, for the mixture of non-Omicron circulating at the time, which is reasonably approximated as Delta in the South African setting).

For the reference scenario, that implies Delta R_0_ is roughly 2.3: the median estimated peak R_t_ since identification of BA.1, 3.4, divided by the median transmissibility multiplier at total immune evasion, 1.5. This may seem remarkably different from the estimates of Delta R_0_ in the 5-7 range (*41*), suggesting that high immune-escape scenarios can be ruled out. However, that interpretation relies on an inaccurate understanding of the previously reported Delta R_0_. That value essentially is derived as: the wild type R_0_ was 2-3 (*42, 43*), Alpha is ∼1.7 times as transmissible as wild type (*39*), Delta ∼1.5 times again as transmissible as Alpha (*44*), and therefore Delta R_0_ is 2-3*1.3*1.7 = 4.4-6.3.

However, that is the Delta R_0_ referenced to the *pre-epidemic period*: that context, and the associated R_0_, no longer applies with the wide variety of changes, particularly in terms of behaviours and associated effective contact rates, that have occurred throughout the pandemic and have not returned to pre-pandemic levels. If we estimate R_0_ for Delta by computing the R_t_ early in the Delta wave in South Africa, determine the necessarily multiplier for transmissibility using the NGM R with estimated proportion susceptible at that time, and then create a counterfactual that removes all immunity and apply that multiplier, we find that the Delta R_c_ at the outset of the Delta wave in South Africa is roughly 2.4 (for the generation interval of approximately 6.3 days assumed in the main analysis).

### Parameters for Next Generation Matrix Calculation

We assume that the relative infectiousness of symptomatic infections is twice that of infections that remain asymptomatic for their entire duration. We use the age-specific relative susceptibility and symptomatic fraction from early COVID-19 pandemic estimates (*34*). We also assume the infectious periods described in that work, which have a mean of 5 days (other distributional features being irrelevant in the next generation method).

**Fig S1.**
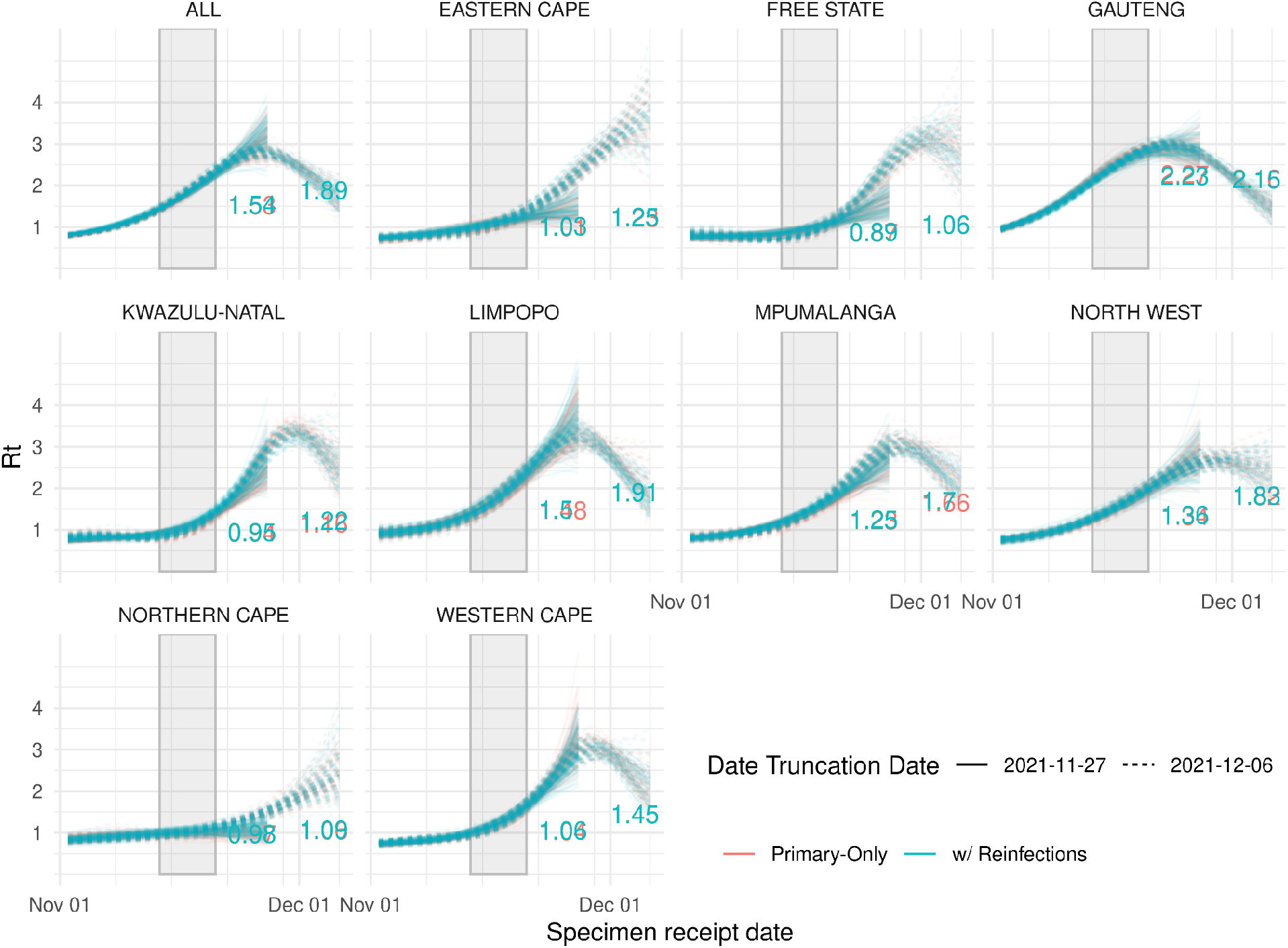
Time-varying reproduction number (R_t_) for all infections versus only primary infections. Using primary infections only versus including reinfections has a very minor effect on estimating R_t_, although changing the cut-off date for the analysis results in noticeably different curves. This effect is minor, however, in the period used for R_t_ ratio estimation (grey box).

### Calculation of vaccine coverage

We obtained weekly numbers of partially and fully vaccinated individuals by age, province, and vaccine product (JnJ or Pfizer) from publicly available data (*45*) and considered vaccine efficacy against observed infection by the Delta variant (*46, 47*) to obtain numbers of effectively vaccinated people in South Africa. To obtain numbers in 5-year age-groups, instead of 10-year age-groups reported by EVDS, we allocated vaccinated individuals from 10-year age-groups to 5-year age-groups according to the corresponding population proportions as estimated by the Thembisa model (*48*). We then calculated vaccination coverage by province and 5-year age-group by dividing the number of effectively vaccinated individuals by the estimated population size. Daily numbers were calculated by linear interpolation. We considered a two-week lag between the day of vaccination and effective protection.

### Calculation of fraction of cases/deaths in the variant era (since December 2021)

We pulled data from “Our World in Data” up to 5^th^ December 2021. We then summed up all cases after first notification of Alpha in the UK (28^th^ September 2021) and all deaths from 28 days later and divide by totals, providing a very rough calculation of the proportion of cases/deaths attributable to variants of concern. For comparison, we also calculated the proportion of all cases (or deaths) in each country from the date (or 28 days later for deaths) that the first incidence of a variant of concern was identified in that country.

|  | Total | Global Total after Alpha (B.1.1.7) in UK | % | Total after first variant per country | % |
| --- | --- | --- | --- | --- | --- |
| Cases | 265,264,696 | 231,862,674 | 87.4% | 146,142,609 | 55.1% |
| Deaths | 5,238,932 | 4,030,271 | 76.9% | 2,664,156 | 50.9% |

### Sensitivity to cut-off date for analysis

To evaluate the sensitivity of our results to the choice of the timeframe used for analysis, we repeated all analyses in the main text using an alternative cut-off specimen receipt date of 27^th^ November 2021, representing the time period before testing levels were affected by the public announcement of detection of the B.1.1.529 lineage (Fig S2).

**Fig S2.**
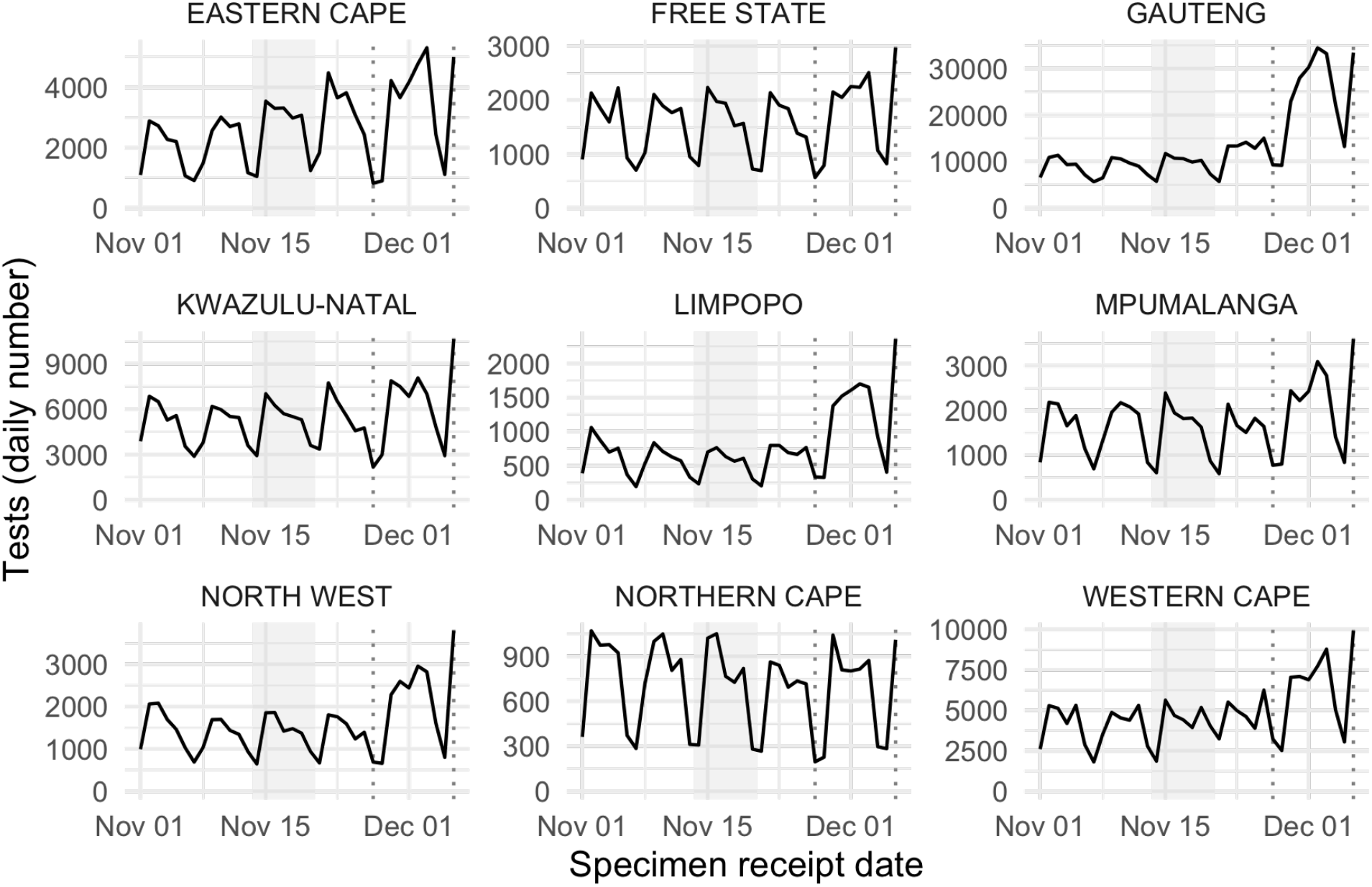
Test volumes by province and specimen receipt date. The solid black curve shows the daily number of tests conducted over time. The light grey area shows the time window used for R_t_ estimation. Dashed vertical lines represent the two cut-off dates used for other analyses: 2021-11-27 (sensitivity analysis) and 2021-12-06 (main analysis). Following the public announcement that a new variant had been identified in South Africa, testing volumes increased substantially, particularly in Gauteng, which was the epicentre of spread of the Omicron variant at that time.

**Table S1:** Fitting parameters for earlier truncation date.

| Province | $\Delta r$ (95% CI) | 50/50 Timing |
| --- | --- | --- |
| Gauteng (GP) | 0.22 (0.20, 0.24) | Nov. 8 (6, 9) |
| KwaZulu-Natal (KZN) | 0.47 (0.33, 0.64) | Nov 19 (17, 20) |

**Fig S3:**
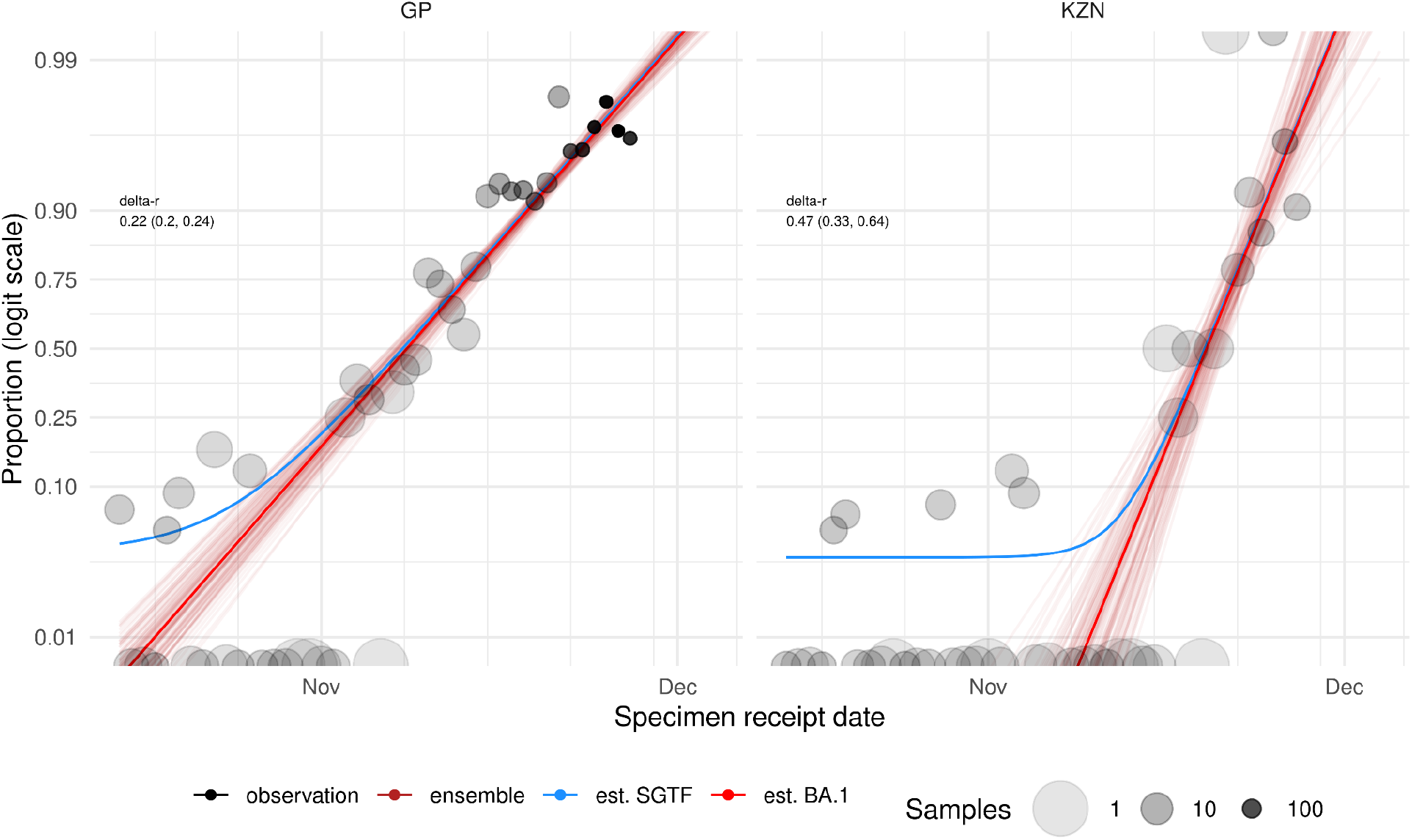
Estimation of the growth advantage of BA.1, using earlier truncation date, for Gauteng and KwaZulu-Natal provinces. The maximum likelihood estimates of BA.1 relative growth rate. Note that the earlier cut off dates limit the provinces for which estimates converge and also tend to lower growth rate estimates. This occurs because earlier fits with less data have much more limited information about the sensitivity of SGTF.

**Fig S4:**
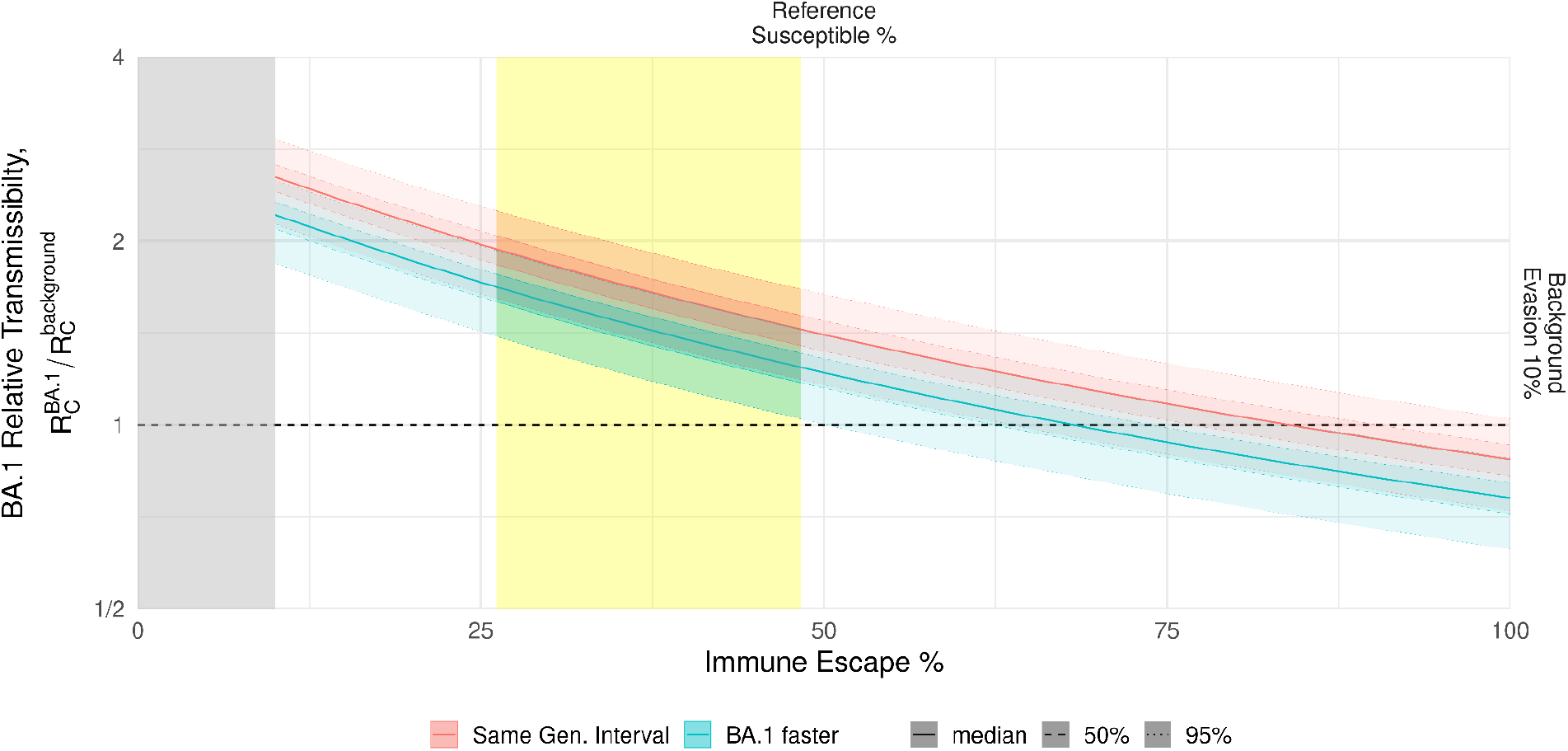
Estimated transmissibility & immune evasion relationship for reference scenario based on estimates for Gauteng, using a cut-off data of 27^th^ November 2021. The red and cyan regions indicate the region of plausibility for relative transmissibility and immune evasion values for BA.1, assuming no change in the generation interval or a shorter generation interval, respectively. The yellow band represents estimated plausible immune escape values, as described in Figure 4 of the main text. The grey band represents values of immune protection that are considered implausible because they would imply greater levels of immune evasion for background variants than for BA.1. The horizontal dashed line indicates equal transmissibility for BA.1 and background variants.

**Fig S5:**
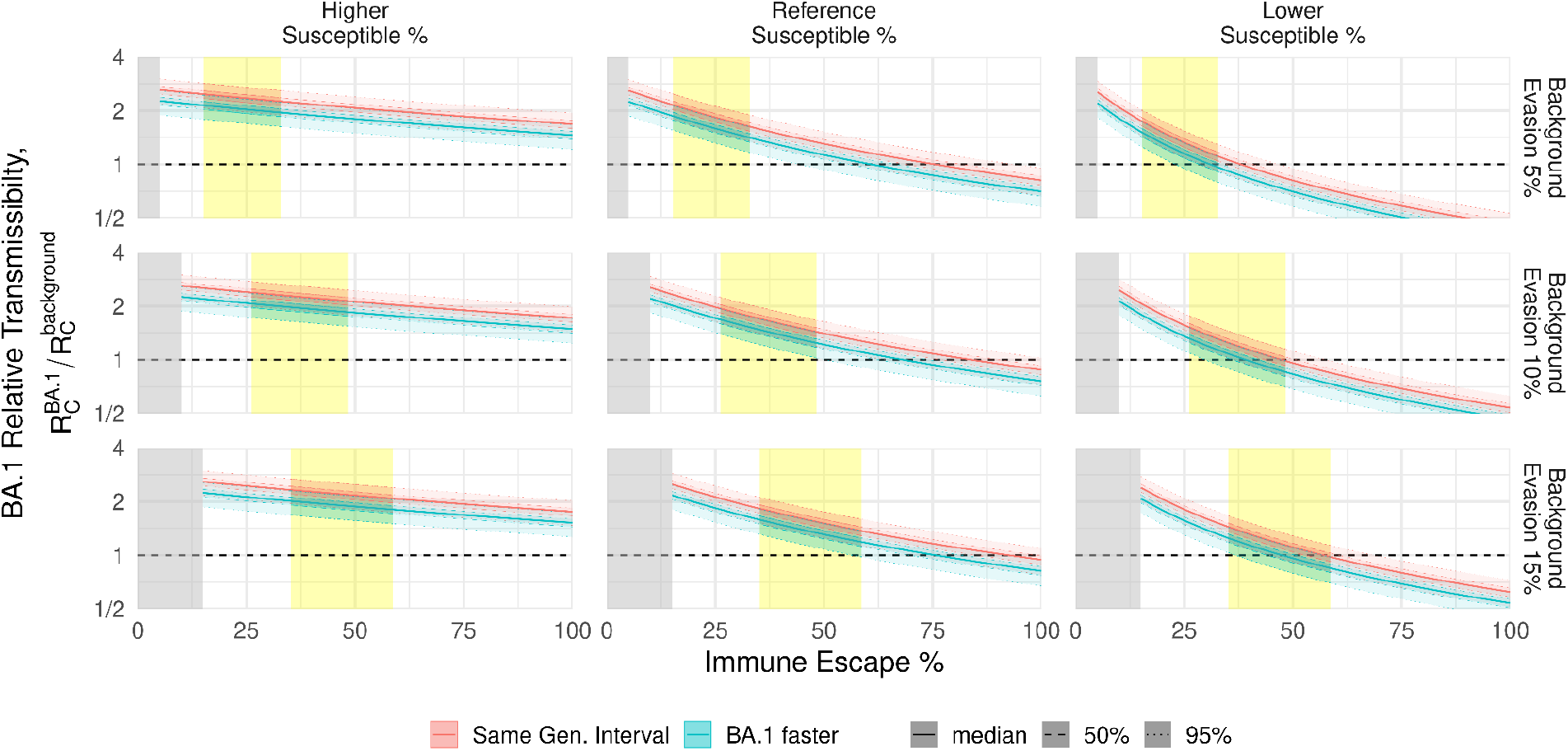
Sensitivity of the plausible transmissibility & immune evasion relationship, using a cut-off data of 27^th^ November 2021. These panels compute the same calculation as in Fig S4, but with varying assumptions regarding the underlying fully susceptible proportion (columns) and the background level of protection provided by prior infection (rows).

**Fig S6:**
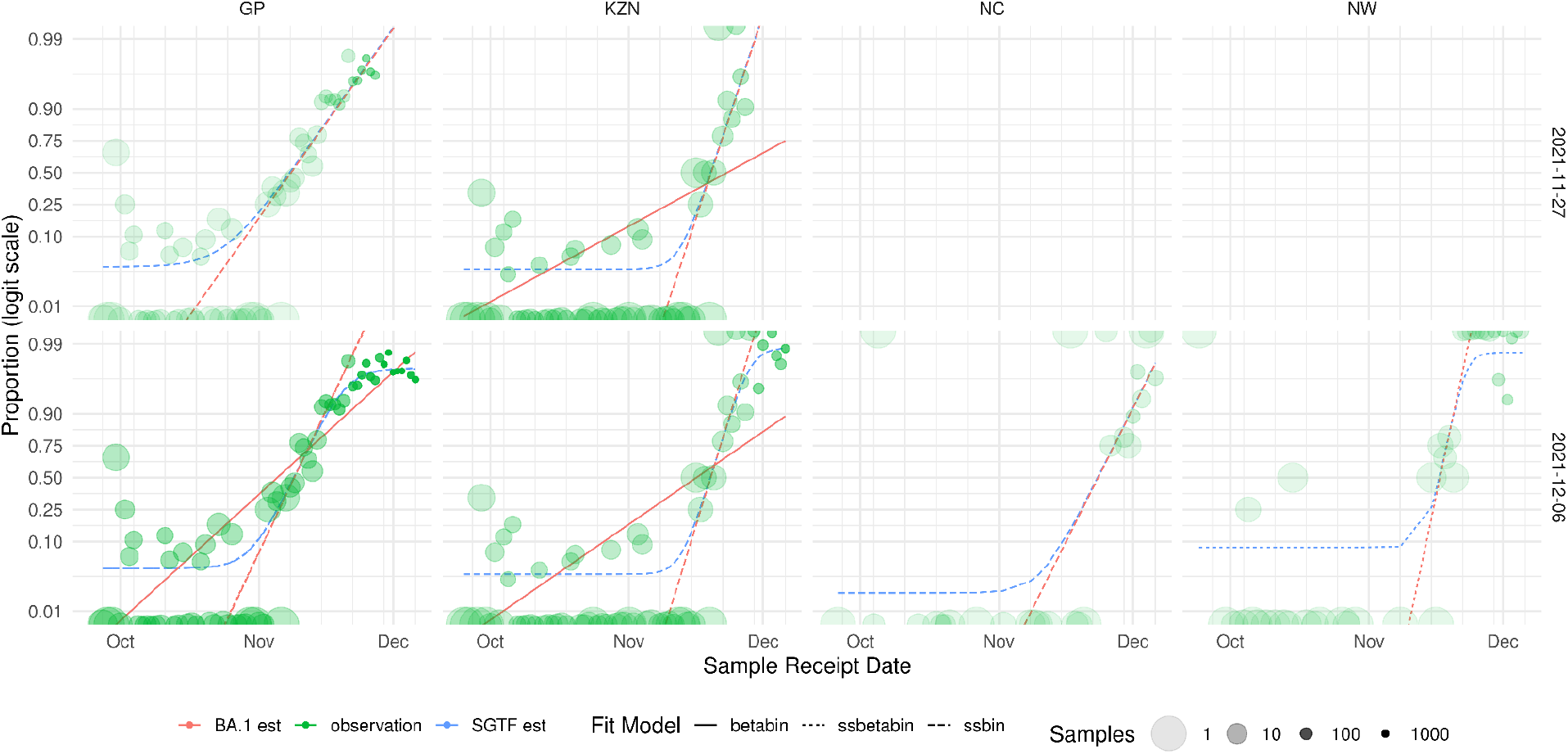
Maximum likelihood fitting comparisons of the different models, by cut-off date. The panels show the different model estimates for different provinces (columns), using different cut off dates for SGTF data (rows). Dashed red and blue lines represent the beta binomial, which accounts for sensitivity and specificity; this is the preferred model for which results are presented in the main text. The solid red lines represent the “straight” beta binomial model (without accounting for sensitivity and specificity), and the dotted lines represent the binomial model with sensitivity and specificity. Only models for which both the estimate and the confidence intervals converge are shown. GP: Gauteng; KZN: KwaZulu-Natal; NC: Northern Cape; NW: North West.

